# Analyzing the effect of relaxing restriction on the COVID-19 outbreak for some US states

**DOI:** 10.1101/2021.04.19.21255759

**Authors:** Mahir Demir, ibrahim Halil Aslan, Suzanne Lenhart

## Abstract

The ongoing pandemic disease COVID-19 has caused worldwide social and financial disruption. As many countries are engaged in designing vaccines, the harmful second and third waves of COVID-19 have already appeared in many countries. To investigate changes in transmission rates and the effect of social distancing in the USA, we formulate a system of ordinary differential equations using data of confirmed cases and deaths in these states: California, Texas, Florida, Georgia, Illinois, Louisiana, Michigan, and Missouri in the USA to be able to investigate changes in transmission rates of the outbreak and effect of social distancing. Our models and the corresponding parameter estimations show social distancing reduces the transmission by 60% to 90%, and thus obeying the movement restriction rules plays a crucial rule to reduce the magnitudes of the outbreak waves. Our analysis shows the current management restrictions do not sufficiently slow the disease propagation.

## 1 Introduction

The virus for COVID-19, SARS-CoV-2, is within the species SARS-like corona viruses. At 125nm, it is marginally bigger than flu and SARS viruses. The closest related virus started from the Rhinolophus bat which is *>* 96% homologous with the current SARS-CoV-2 virus and it is only 79% homologous with the original SARS CoV [1].

Clearly, transmission occurs human-to-human and from close face-to-face social contact, particularly in small enclosed spaces. The transmission can be enormous in a brief period of time with thousands of new cases every day. While the symptoms of COVID-19 can range broadly from asymptomatic illness to pneumonia and life-threatening complications, the most common symptoms are fever, dry cough, fatigue, nasal congestion, sore throat and diarrhoea. Elderly patients with preexisting respiratory or cardio-vascular conditions are the greatest risk group for extreme complications [1, 2, 3, 4].

Since late January, 2020, major public health interventions have been implemented over China and other countries to slow down the spread of the disease. For example, strong social separating measures and mobility limitations were facilitated and executed by the central and local governments in many countries. These huge social restrictions and alternative interventions have tremendous costs, in terms of both direct costs and loss of financial opportunities [5].

The European Commission’s roadmap has warned people “the way back to normality will be very long and masks, gloves, tests will become usual until a vaccine available,” while a widespread vaccine will not be accessible for almost one year [6]. Relaxing the interventions when the epidemic size was small would increase the cumulative number of cases and lead to the second wave [7, 1, 5]. Dr. Anthony Fauci said the second wave is inevitable and if states begin removing the restrictions too early, the nation could seen a rebound of the virus that would “get us right back in the same boat [8, 9].” The UK Prime Minister pointed out if the UK eases the restriction too rapidly, all the exertion and sacrifice would be thrown away [10]. Despite of all these concerns, some states in the USA move to remove some of the restrictions, and some European nations with the most cases of COVID-19-related deaths are starting to lift their lockdowns and have begun facilitating some schools and businesses to reopen. The countries such as Denmark, Austria and Czech Republic are moving back to a sense of normality but a new kind [8, 9, 10, 6].

There are several studies recently published related to COVID-19 and its effect of social distancing. One of them investigated the effect of the lockdowns for some countries in Europe and in the USA by utilizing an SEIR system of ordinary differential equations (ODEs) [11]. This study compares the basic reproduction number before and after lockdowns. Another work using a SEIR ODE model discussed the timing and corresponding effects of social distancing as control measures on the burden of COVID-19 in USA [12]. Similarly, a study quantified the effectiveness of social distancing and discussed the relaxation of the social distancing without causing a second wave in Ontario, Canada [13]. However, we have not found any study analyzing the effect of relaxing restrictions by using a SIR type mathematical models for some specific states in the USA.

In this study, we use mathematical models to examine the effect of relaxing restrictions on the cumulative number of confirmed cases and deaths. We start with introducing the mathematical models in the following section and then discuss fitting the parameters to the data that are retrieved by The New York Times [14]. Finally, the numerical simulation and the behavior of propagation of the disease with relaxing restrictions are discussed in the final section.

## 2 Models

We utilize a compartmental ODE model proposed by Aslan et al. in [15] and modify it to include relaxing the restrictions. The ODE system from [15] is denoted by Model 1, and the modified ODE system is Model 2. These two models are used to investigate and project the COVID-19 spread in some states of USA. Model 1 is the preliminary model with ordinary differential equations (ODEs) with the rates of change among the compartments of the population given in Figure 1. This model will represent the population compartments before the relaxing restrictions start. Model 2 is a modified version of Model 1 given in Figure 2 will present the compartments after relaxing restrictions The details of these models are given in following subsections.

**Figure 1:**
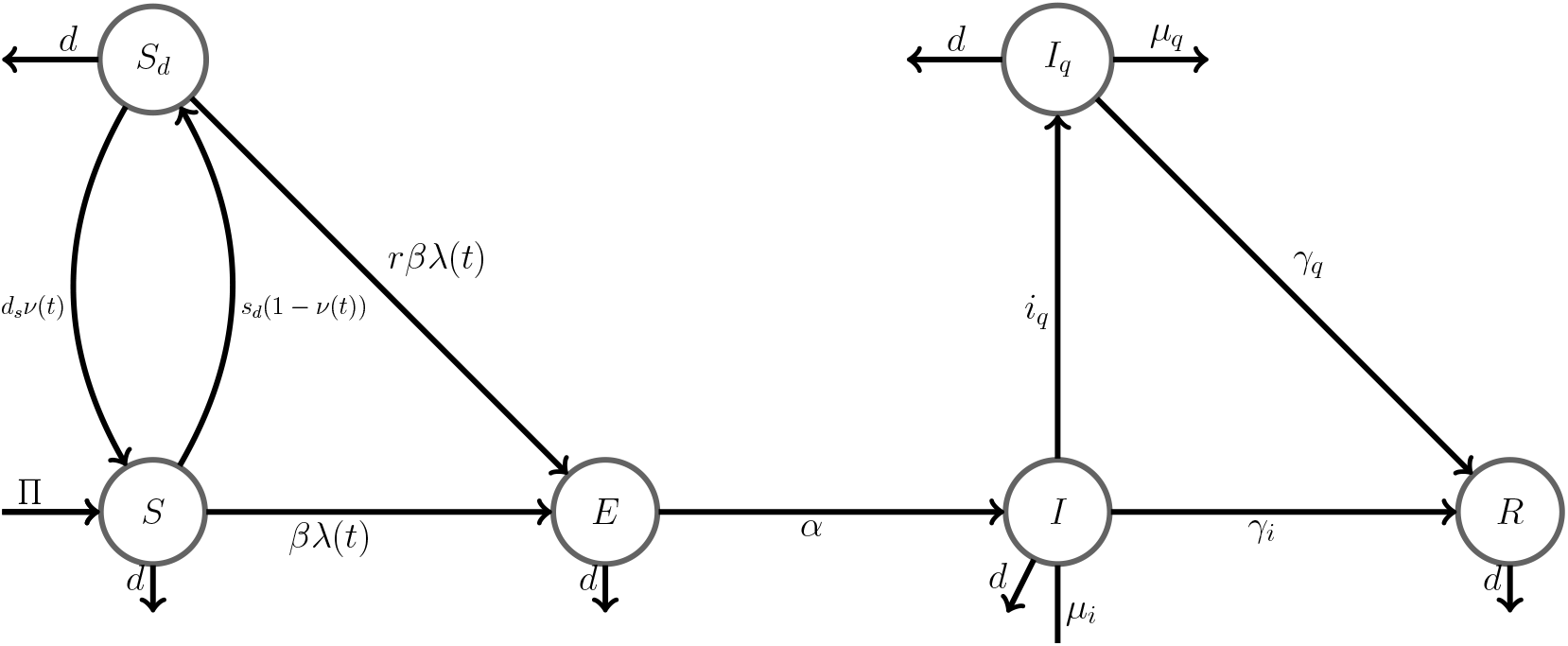
Flow diagram illustrating the disease transitions among the compartments

**Figure 2:**
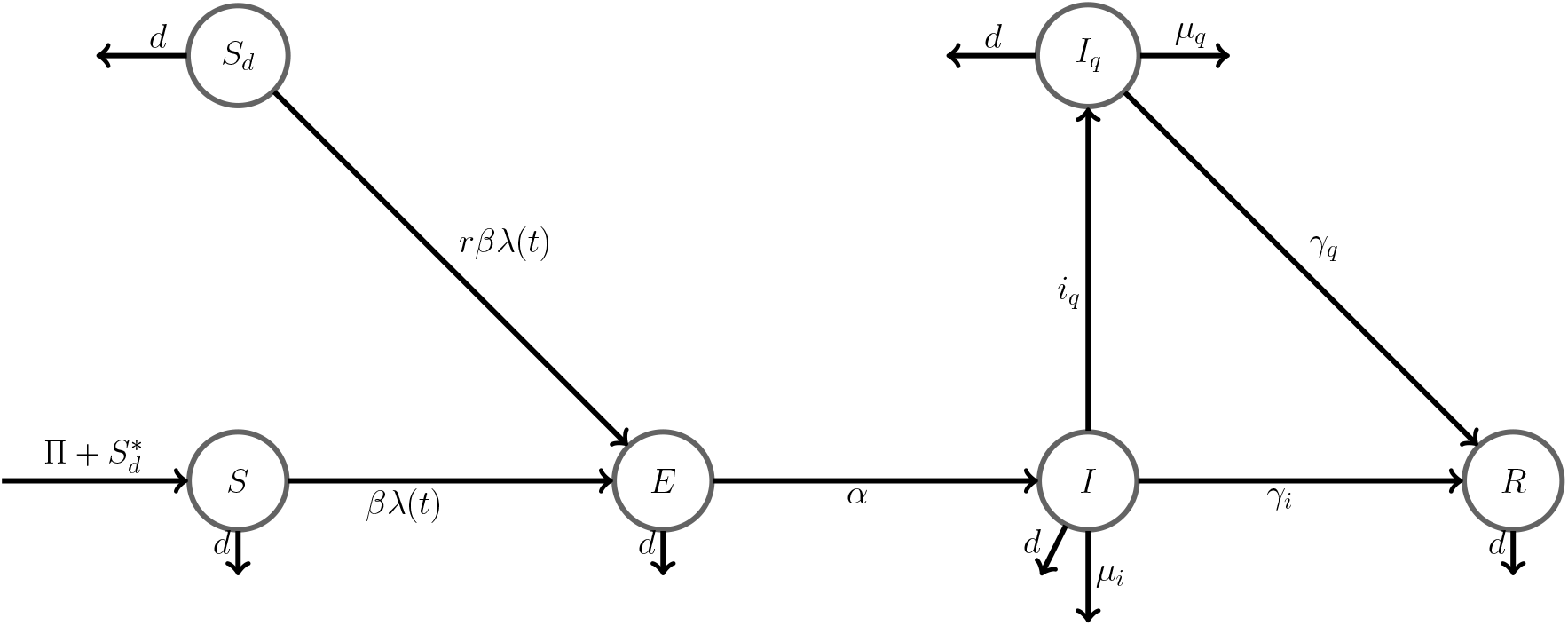
Flow diagram illustrating the disease transitions among the compartments with impulse action

### 2.1 Model 1 (the preliminary model)

Model 1 is simulated from the date of first case appears to the date of first relaxing restrictions and employs a type of feedback mechanism. The rate of confirmed cases (i.e., the number of individuals who are sick, test positive, and are subsequently isolated per day) affects the rate of susceptible individuals moving into social distancing. When the number of confirmed cases increases, the number of people in social distancing will increase. However, when the daily number of confirmed new cases reaches a low level, social distancing mandates are relaxed, allowing population members to move back into the susceptible compartment with a higher transmission rate. The model simulations illustrate that this decrease in social distancing could result in a new wave of outbreak.

In this model, the Susceptible compartment *S* represents individuals in the population who have not yet been exposed to COVID-19, and are not actively practicing social distancing, and *S*_*d*_ individuals are susceptible who are social distancing in the population. These individuals are strictly practicing the social distancing rules such as using masks, keeping 6-foot distance. Individuals in the Exposed compartment *E* have been exposed to COVID-19 but are not yet able to transmit the disease, while those in the Infected *I* compartment are asymptomatic or only exhibiting mild symptoms but have not been tested, confirmed positive, nor consequently isolated. Individuals in the isolated compartment *I*_*q*_ have tested positive and have been isolated, and finally the individuals in the recovered compartment R *R* individuals had been infected and have recovered. Figure 1 shows the flow of people among these compartments.

Where

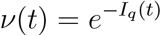

represents the product function of the transition rate to social distancing class. Note that, as the number of new cases increases the value of *ν*(*t*) decreases and the number of individuals transitioning to social distancing class increases. The force of infection is

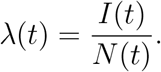

Based on these compartments according to the disease progression, the following system of ordinary differential equations (ODEs) represents the flow among these compartments.

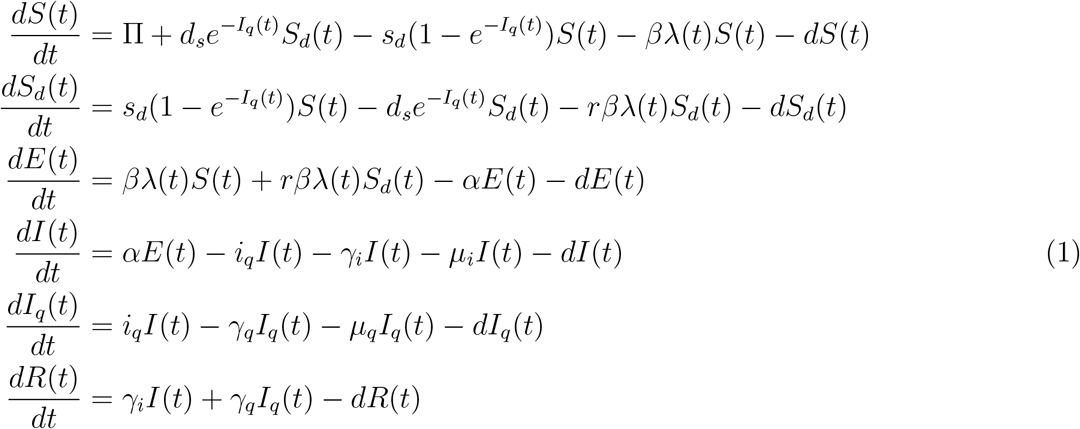

with *S*(0) > 0, *S*_*d*_(0) ≥ 0,*E*(0) ≥ 0, *I*(0) ≥ 0, *I*_*q*_(0) ≥ 0, *R*(0) ≥ 0 and

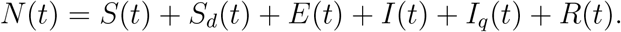

The description of parameters is given in Table 1.

**Table 1:** Description of model parameters

| Parameter | Description | Unit |
| --- | --- | --- |
| $\beta$ | Disease transmission rate | $\frac{1}{\text{day}}$ |
| $d_s$ | Rate of release from social distancing compartment | $\frac{1}{\text{day}}$ |
| $s_d$ | Transition rate to social distancing compartment | $\frac{1}{\text{day}}$ |
| $\alpha$ | Rate of exposed individuals becoming infected | $\frac{1}{\text{day}}$ |
| $\gamma_i$ | Recovery rate due to natural immune response | $\frac{1}{\text{day}}$ |
| $\gamma_q$ | Recovery rate due to hospitalization | $\frac{1}{\text{day}}$ |
| $\mu_i$ | Death rate of disease in infectious compartment | $\frac{1}{\text{day}}$ |
| $\mu_q$ | Death rate due to disease in isolated infectious compartment | $\frac{1}{\text{day}}$ |
| $i_q$ | Transition rate to isolated infectious compartment | $\frac{1}{\text{day}}$ |
| $r$ | Reduction rate due to social distancing | - |
| $\Pi$ | Recruitment rate | $\frac{\text{Individual}}{\text{day}}$ |
| $d$ | Natural death rate | $\frac{1}{\text{day}}$ |

To accurately capture the local outbreak dynamics in each state, the key parameters of the model were estimated for each state separately by using data available in [14, 16] from the date of first case appears to the date of the first relaxing restrictions.

### 2.2 Model 2

Model 2 starts playing role with relaxing the restrictions. Based on the decision of government authorities and hdealth agencies, a proportion of the people in *S*_*d*_ compartment moves to *S* compartment with an impulse action on the date of the first relaxing restrictions, and the system (1) switch to a new dynamical system given in (2). The initial conditions of this new system are the values of system (1) at final time, which corresponds the first day of relaxing restrictions. Figure 2 shows the flow of people among the compartments for the second model.

This second model differs from the previous model with two ways. The first difference is the impulse action which moves a proportion of the susceptible population in the social distancing compartment, *S*_*d*_, to the susceptible compartment, *S*. The second difference is that there is no transition between susceptible compartment and the social distancing compartment. The dynamical system representing this flow is given by a system of ODEs as following

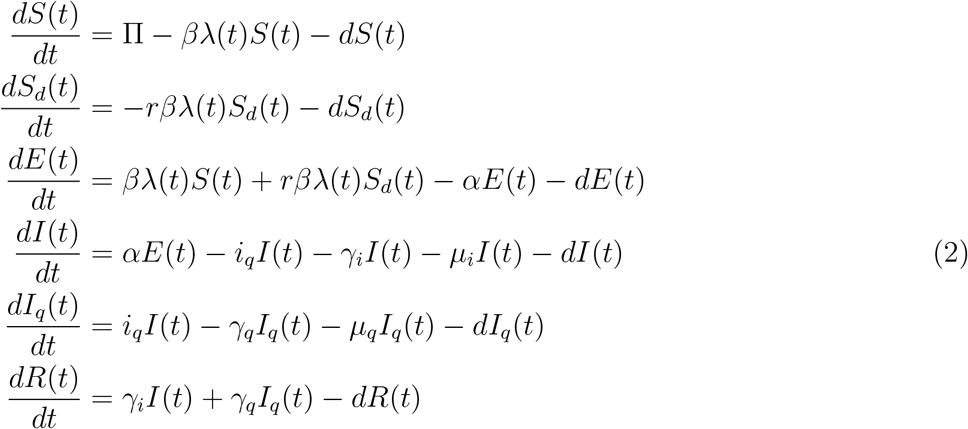

with *S*(*T* ^+^) = *pS*_*d*_(*T* ^−^) + *S*(*T* ^−^), *S*_*d*_(*T* ^+^) = (1 − *p*)*S*_*d*_(*T* ^−^), *E*(*T* ^+^) = *E*(*T* ^−^), *I*(*T* ^+^) = *I*(*T* ^−^), *I*_*q*_(*T* ^+^) = *I*_*q*_(*T* ^−^), *R*(*T* ^+^) = *R*(*T* ^−^) where *T* is the date of relaxing restrictions and *pS*_*q*_(*T* ^−^) is the proportion of the people in the social distancing compartment move to susceptible compartment with impulse action. This model is fitted to the data multiple times from the first date of relaxing restriction to March 3th, 2021 and then the model is used to forecast for 15 days. In the fitting processes, the parameters,*β, r, µ*_*q*_, *µ*_*i*_, *i*_*q*_, *γ*_*q*_ are estimated.

## 3 Parameter estimation and adapting parameter values

We estimate the baseline parameters in the systems (1) and (2) by fitting our models with cumulative number of cases and deaths data provided by [14] and [16]. We use the Ordinary Least Squares (OLS) method and minimize the sum of the squares of differences between the daily reported data and those predicted by our model. Given a parameter set, the corresponding goodness of fit is measured by computing the associated relative error using the formula

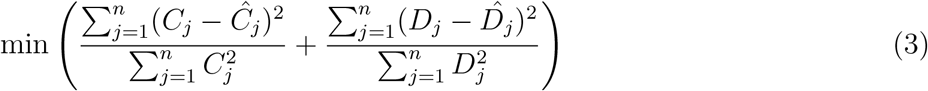

where *C*_*j*_ and *Ĉ*_*j*_ are respectively the reported (from data) and simulated cumulative confirmed (infected) cases, and *D*_*j*_ and 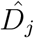 are respectively the reported and simulated cumulative deaths, with *j* = 1, 2, …, *n* as the index of the daily reported and simulated cumulative confirmed number of cases and deaths. In estimation of the cumulative number of COVID-19 cases, we consider cumulative number of confirmed cases in *I*_*q*_ compartment. In estimation of the number of COVID-19 deaths, we sum the number of deaths coming from the infected compartment *I* and the confirmed (infected) compartment *I*_*q*_. Note that the natural deaths in the infected compartment *I* and the confirmed (infected) compartment *I*_*q*_ are also included in the total number of deaths. We used an ode45 solver with fmincon and multistart by using the Optimization Toolbox of MATLAB with 10 points and used the initial conditions of each state in USA.

We estimated the following parameters for each state: disease transmission rate, *β*, reduction rate due to social distancing, *r*, transition rate to isolated infected compartment, *i*_*q*_, transition rate to social distancing compartment, *s*_*d*_, rate of release from social distancing compartment, *d*_*s*_, recovery rate due to hospitalization, *γ*_*q*_, death rate due to disease in isolated infectious compartment, *µ*_*q*_, and death rate due to disease in infectious compartment, *µ*_*i*_. We estimated these parameters for each state separately by using state specific data. We used the same natural death rate *d* = 2.3 * 10^−5^ and recruitment rate Π depending of population of each state provided by the WHO[17]. For each state, we used 4.2 days for the average incubation period, 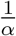 from [18, 19, 20], so *α* = 0.2381. Finally, we estimated recovery rate due to natural immune response, *γ*_*i*_ = 0.111 and by using the total number of cases and deaths in the whole USA and used these estimated values for each state. Thus, we did not estimate *γ*_*i*_ for each state separately since we assumed these values do not change from state to state in the USA.

## 4 Numerical Simulations

In this section, we discuss the forecasting and fit of models for California, Texas, Florida, Georgia, Illinois, Louisiana, and Michigan. We fitted the models to the data for each state separately, then first investigated the relaxing restrictions and their effect in propagation of COVID-19 for these states. After that we forecasted the cumulative number of deaths, the cumulative number of cases, and daily cases with different scenarios by changing the social interaction among the people.

To be able to investigate relaxing restrictions in these states, we fitted the Model 1 to data from the date of first case appears to the date of relaxing restriction start, then the model 2 given in (2) is fitted to the data multiple times depends on the date of relaxing restriction and behavior of data. The date of relaxing restrictions for each state is adjusted according to the government resources and the data [21]. After estimating the parameters with these model fits, we investigated these parameters for each state to indicate which relaxing restrictions were better to contain or slow down the spread of COVID-19.

Since we also would like to predict trajectories of the COVID-19 outbreak in these states, we consider three scenarios until March 18, 2021 in this study. Scenario 1 is shown with green line **−** where we forecast with the parameters that are estimated in the currently active restrictions. Scenario 2 is shown with orange line **−** where we forecast with the parameters that are estimated in the currently active restrictions but increase the transmission by 50%. Scenario 3 is shown with light green line **− *−*** where we forecast with the parameters that are estimated in the currently active restrictions but decrease the transmission by 50%. These three scenarios may occur in any states.

### 4.1 California

According to the time series of confirmed cases and deaths from March 2020 to March 2021, the second and particularly third waves are much bigger than the first wave (see Figure 3). Based on the estimated parameters of our model, we also see the larger transmission rates as the second and third waves start (see Table 2). We do not see any significant changes in social distancing levels due to no change in reduction rate, *r* (see Table 2). This indicates that this social distancing level is not enough to completely contain the outbreak in California. We also see the smallest transmission rate, *β* between January 10, 2021 and March 2, 2021.

**Table 2:** Transmission rate, *β*, the reduction rate due to the social distancing, *r*, and the reduced transmission rate due to the social distancing, *rβ* for each corresponding fit in the state of California

| Fits | $\beta$ | $r$ | $r\beta$ | Date |
| --- | --- | --- | --- | --- |
| First fit | 0.86 | 0.1 | 0.086 | January 25, 2020- May 19, 2020 |
| Second fit | 0.91 | 0.15 | 0.1365 | May 20, 2020- July 9, 2020 |
| Third fit | 0.66 | 0.15 | 0.099 | July 10, 2020- October 9, 2020 |
| Fourth fit | 1.32 | 0.15 | 0.198 | October 10, 2020- January 9, 2021 |
| Fifth fit | 0.51 | 0.4 | 0.204 | January 10, 2021- March 2, 2021 |

**Figure 3:**
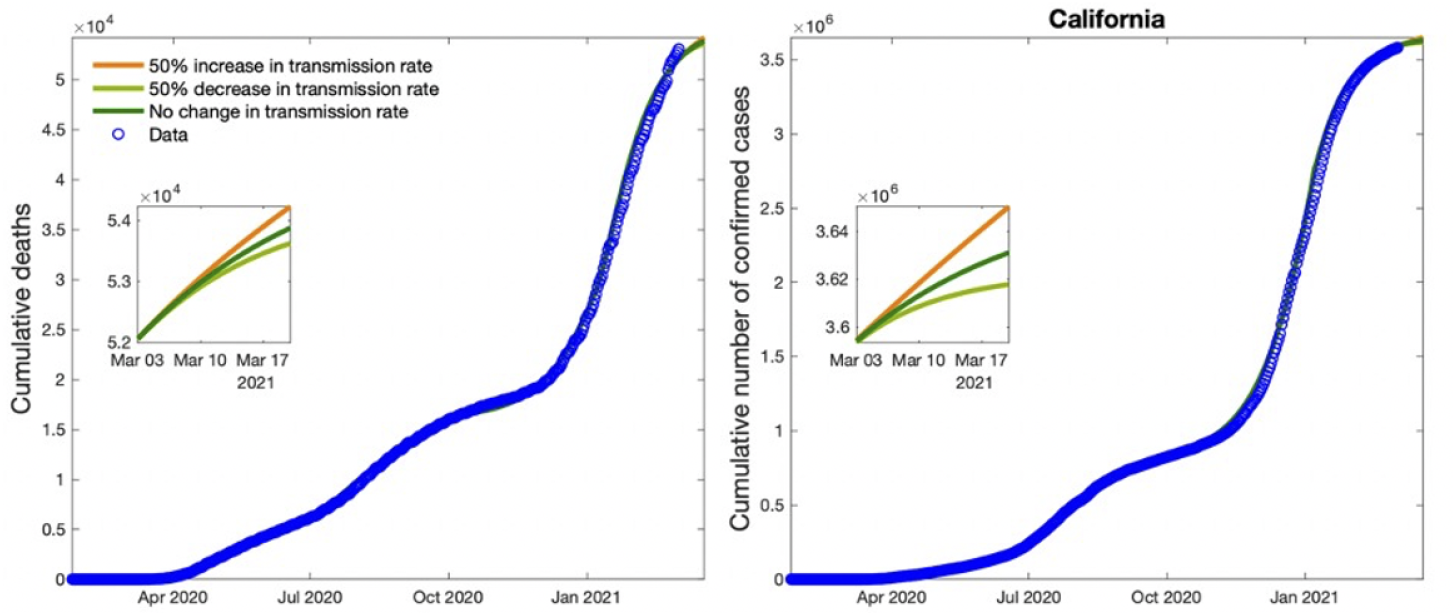
Plots show cumulative cases, deaths, and new cases in California with data. The small boxes in the figures refer to fifteen days forecasting by varying the transmission rate *β*, 50% above and below its estimated value.

We also forecast the cumulative number of cases and deaths from March 2, 2021 to March 25, 2021. Depending on our 3 scenarios, we forecasted possible number of cases and deaths for California (see Table 3).

**Table 3:** Forecasting of the total number of cases, deaths and case fatality rate (CFR) with forecasting up to February 15^th^, 2021 in California, USA

| California | Total # of Cases | Total # of Deaths | CFR |
| --- | --- | --- | --- |
| Scenario 1 | 3,631,120 | 53,881 | 0.015 |
| Scenario 2 | 3,650,290 | 54,231 | 0.015 |
| Scenario 3 | 3,617,830 | 53,628 | 0.015 |

### 4.2 Texas

As it can be seen in Figure 4, the second and third waves are larger than the first wave. Based on our estimated parameters, we see larger transmission rates in the second and third waves which are causing important increases in confirmed cases and deaths in Texas. These increases in transmission rates might be due to decreases in social distancing during the second and third waves (see Table 4). Similar to California, we see the smallest transmission rate between January 10, 2021 and March 2, 2021. We also forecast the cumulative number of cases and deaths from March 2, 2021 to March 25, 2021. Depending on our 3 scenarios, we forecasted possible number of cases and deaths for Texas (see Table 5).

**Table 4:** Transmission rate, *β*, the reduction rate due to the social distancing, *r*, and the reduced transmission rate due to the social distancing, *rβ* for each corresponding fit in the state of Texas

| Fits | $\beta$ | $r$ | $\beta r$ | Date |
| --- | --- | --- | --- | --- |
| First fit | 1.13 | 0.12 | 0.1356 | January 25, 2020- May 19, 2020 |
| Second fit | 1.31 | 0.14 | 0.1834 | May 20, 2020- July 9, 2020 |
| Third fit | 0.92 | 0.15 | 0.138 | July 10, 2020- September 11, 2020 |
| Fourth fit | 1.28 | 0.18 | 0.2304 | September 12, 2020- January 22, 2021 |
| Fifth fit | 0.84 | 0.24 | 0.2016 | January 23, 2021- March 2, 2021 |

**Table 5:** Forecasting of the total number of cases, deaths and case fatality rate (CFR) with forecasting up to February 15^th^, 2021 in Texas, USA

| California | Total # of Cases | Total # of Deaths | CFR |
| --- | --- | --- | --- |
| Scenario 1 | 2,778,140 | 45,858 | 0.016 |
| Scenario 2 | 2,867,040 | 46,805 | 0.016 |
| Scenario 3 | 2,732,660 | 45,316 | 0.016 |

**Figure 4:**
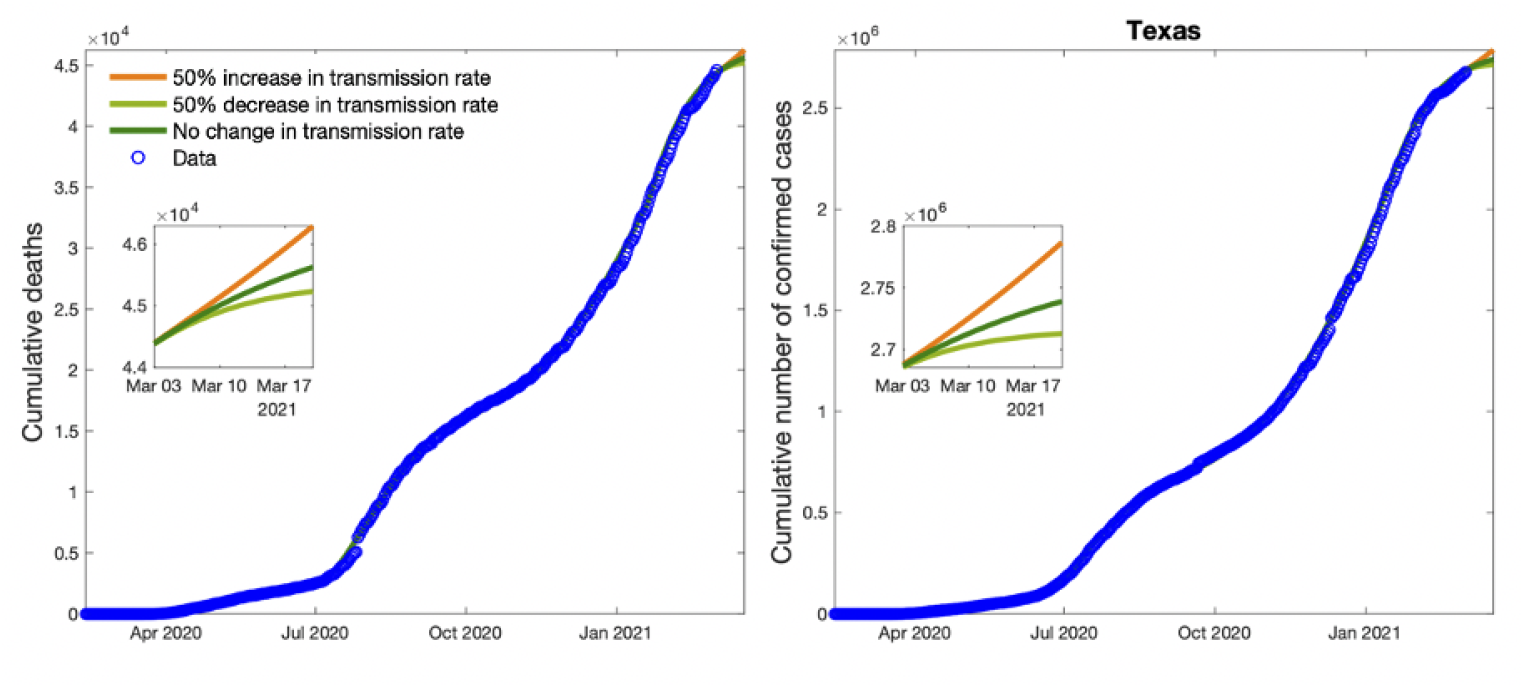
Plots show cumulative cases, deaths, and new cases in Texas with data. The small boxes in the figures refer to fifteen days forecasting by varying the transmission rate *β* 50% above and below its estimated value.

### 4.3 Florida

Similar to Texas, the second and third waves are again larger than the first wave in Florida (see Figure 5). Depending on our estimated parameters, we see the larger transmission rates in the second and third waves in Florida (see Table 6). These increases in transmission rates might be due to decreases in social distancing during the second and third waves (see Table 6) in Florida. We also forecast the cumulative number of cases and deaths from March 2, 2021 to March 25, 2021. Depending on our 3 scenarios, we forecasted possible number of cases and deaths for Florida (see Table 7).

**Table 6:** Transmission rate, *β*, the reduction rate due to the social distancing, *r*, and the reduced transmission rate due to the social distancing, *rβ* for each corresponding fit in the state of Florida

| Fits | $\beta$ | $r$ | $r\beta$ | Date |
| --- | --- | --- | --- | --- |
| First fit | 1.65 | 0.13 | 0.2145 | January 25, 2020- May 19, 2020 |
| Second fit | 2.14 | 0.15 | 0.321 | May 20, 2020- July 9, 2020 |
| Third fit | 1.04 | 0.15 | 0.156 | July 10, 2020- September 9, 2020 |
| Fourth fit | 1.78 | 0.17 | 0.3026 | September 10, 2020- January 9, 2021 |
| Fifth fit | 1.31 | 0.21 | 0.2751 | January 10, 2021- March 2, 2021 |

**Table 7:** Forecasting of the total number of cases, deaths and case fatality rate (CFR) with forecasting up to February 15^th^, 2021 in Florida, USA

| California | Total # of Cases | Total # of Deaths | CFR |
| --- | --- | --- | --- |
| Scenario 1 | 2,016,000 | 32,996 | 0.016 |
| Scenario 2 | 2,042,620 | 33,180 | 0.016 |
| Scenario 3 | 1,974,930 | 32,681 | 0.016 |

**Figure 5:**
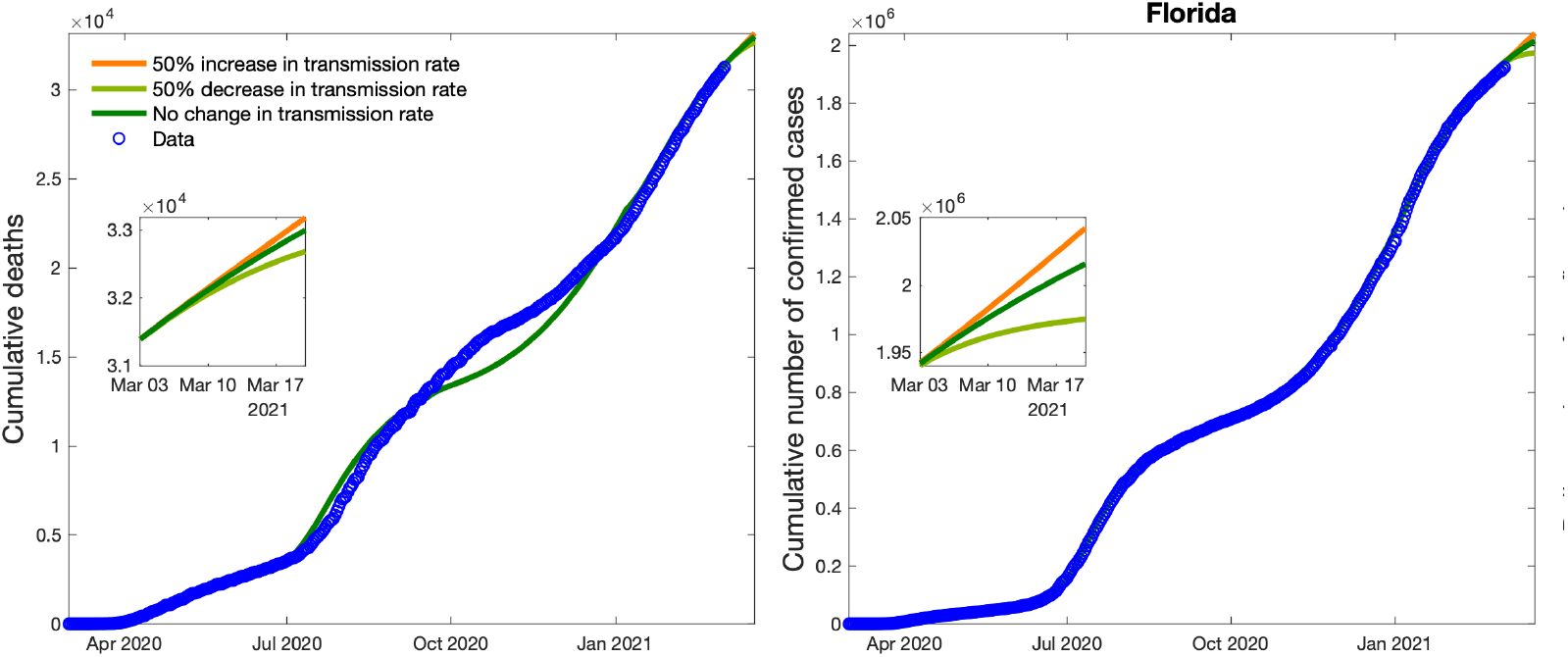
Plots show cumulative cases, deaths, and new cases in Florida with data. The small boxes in the figures refer to fifteen days forecasting by varying the transmission rate *β* 50% above and below its estimated value.

### 4.4 Georgia

According to the time series of confirmed cases and deaths from March 2020 to March 2021, the second and particularly third waves are bigger than the first wave (see Figure 6). Based on the estimated parameters of our model, we see larger transmission rates during the first wave, the second and third waves (see Table 8). Even if we see the largest transmission rate in the first wave, the social distancing level is 0.1, which causes 90 percent reduction in transmission rate.

**Table 8:**
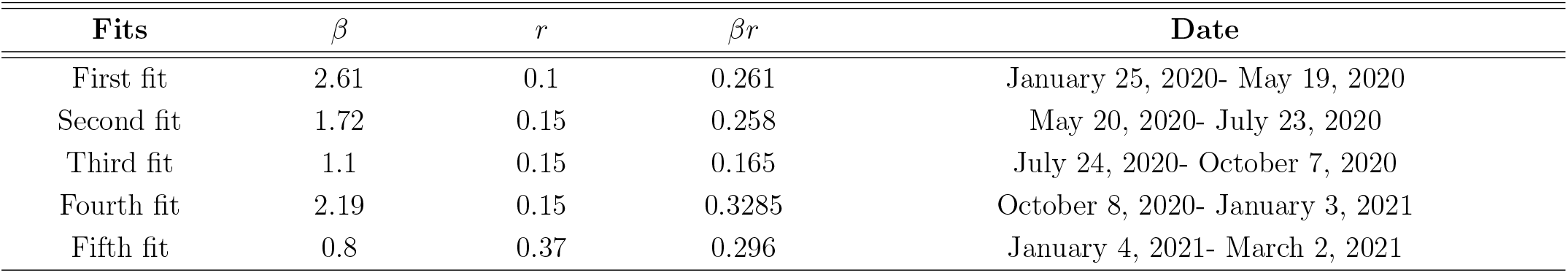
Transmission rate, *β*, the reduction rate due to the social distancing, *r*, and the reduced transmission rate due to the social distancing, *rβ* for each corresponding fit in the state of Georgia

**Figure 6:**
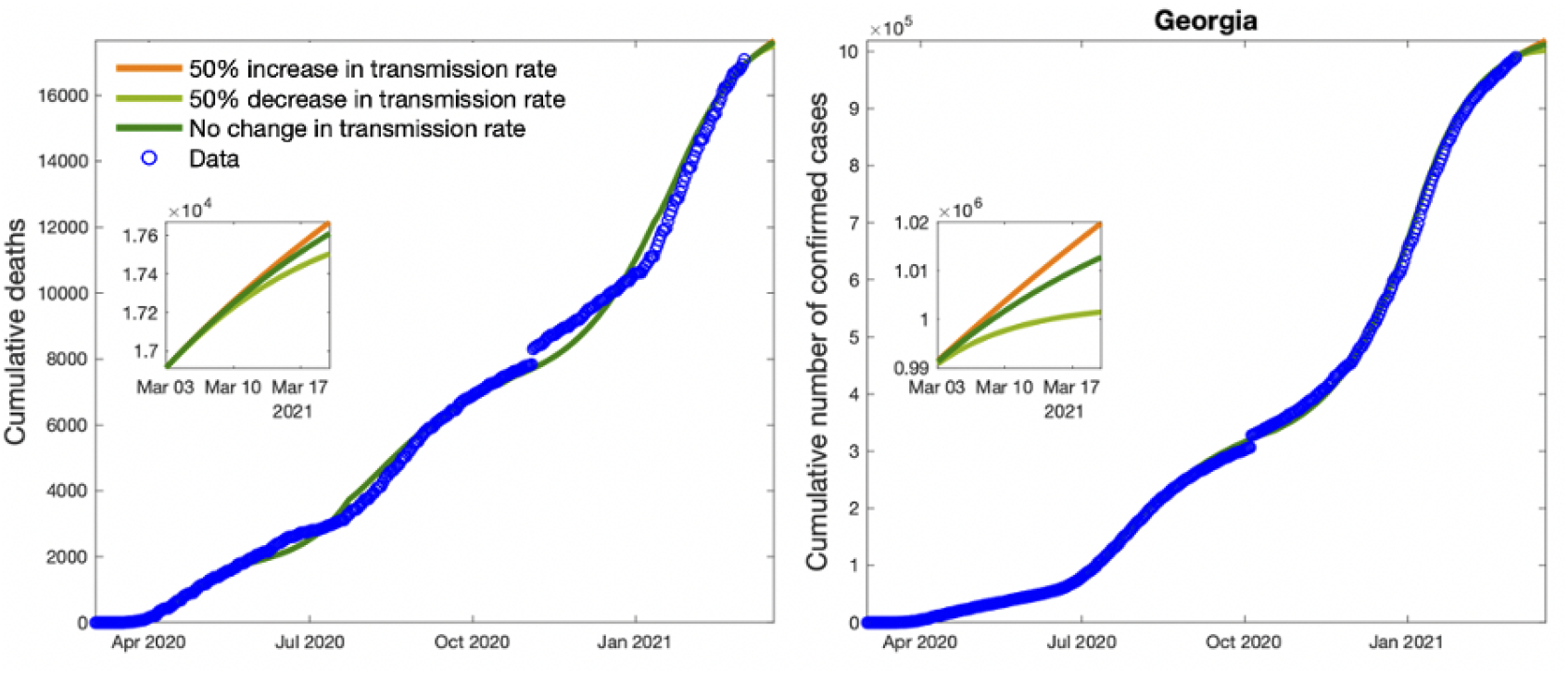
Plots show cumulative cases, deaths, and new cases in Georgia with data. The small boxes in the figures refer to fifteen days forecasting by varying the transmission rate *β* 50% above and below its estimated value.

We do not see any significant changes in social distancing levels in the second and third waves since we have no changes in reduction rate, *r* (see Table 9). This indicates that this social distancing level is not enough to completely contain the outbreak in Georgia in the second and third waves. We also see the smallest transmission rate, *β* between January 10, 2021 and March 2, 2021 and the largest transmission rate, *β* between January 25, 2020 and May 19, 2020. We also forecast the cumulative number of cases and deaths from March 2, 2021 to March 25, 2021. Depending on our 3 scenarios, we forecasted possible number of cases and deaths for California (see Table 9).

**Table 9:** Forecasting of the total number of cases, deaths and case fatality rate (CFR) with forecasting up to February 15^th^, 2021 in Georgia, USA

| **California** | **Total # of Cases** | **Total # of Deaths** | **CFR** |
| --- | --- | --- | --- |
| Scenario 1 | 1,006,840 | 17,521 | 0.017 |
| Scenario 2 | 1,013,860 | 17,580 | 0.017 |
| Scenario 3 | 995,569 | 17,417 | 0.017 |

### 4.5 Illinois

As it can be seen in Figure 7, we have 3 waves and before the second wave ended the third wave occurred. Based on our estimated parameters, we see larger transmission rates in the first, second and third waves which are causing important increases in confirmed cases and deaths in Illinois. These large transmission rates in second and third waves might be due to decreases in social distancing during the second and third waves (see Table 10). Even if we see the largest transmission rate in the first wave, the social distancing level is 0.1, which causes 90 percent reduction in transmission rate.

**Table 10:**
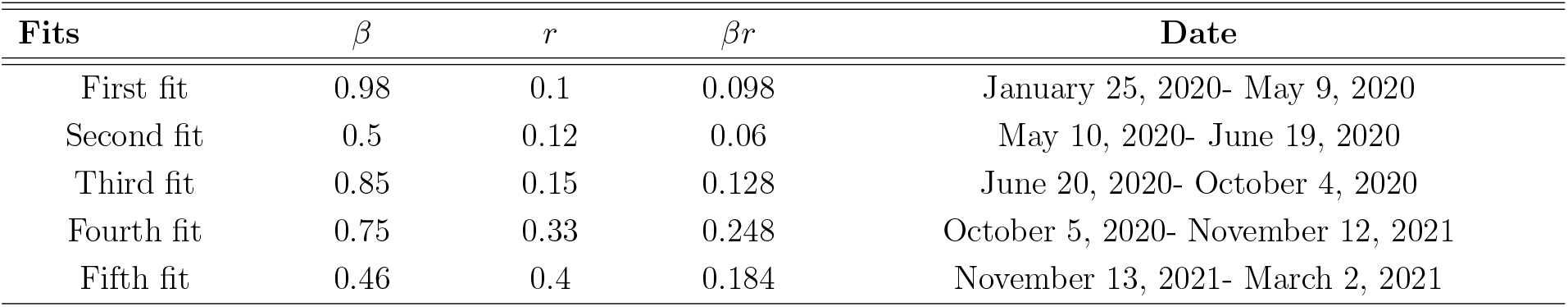
Transmission rate, *β*, the reduction rate due to the social distancing, *r*, and the reduced transmission rate due to the social distancing, *rβ* for each corresponding fit in the state of Illinois

**Figure 7:**
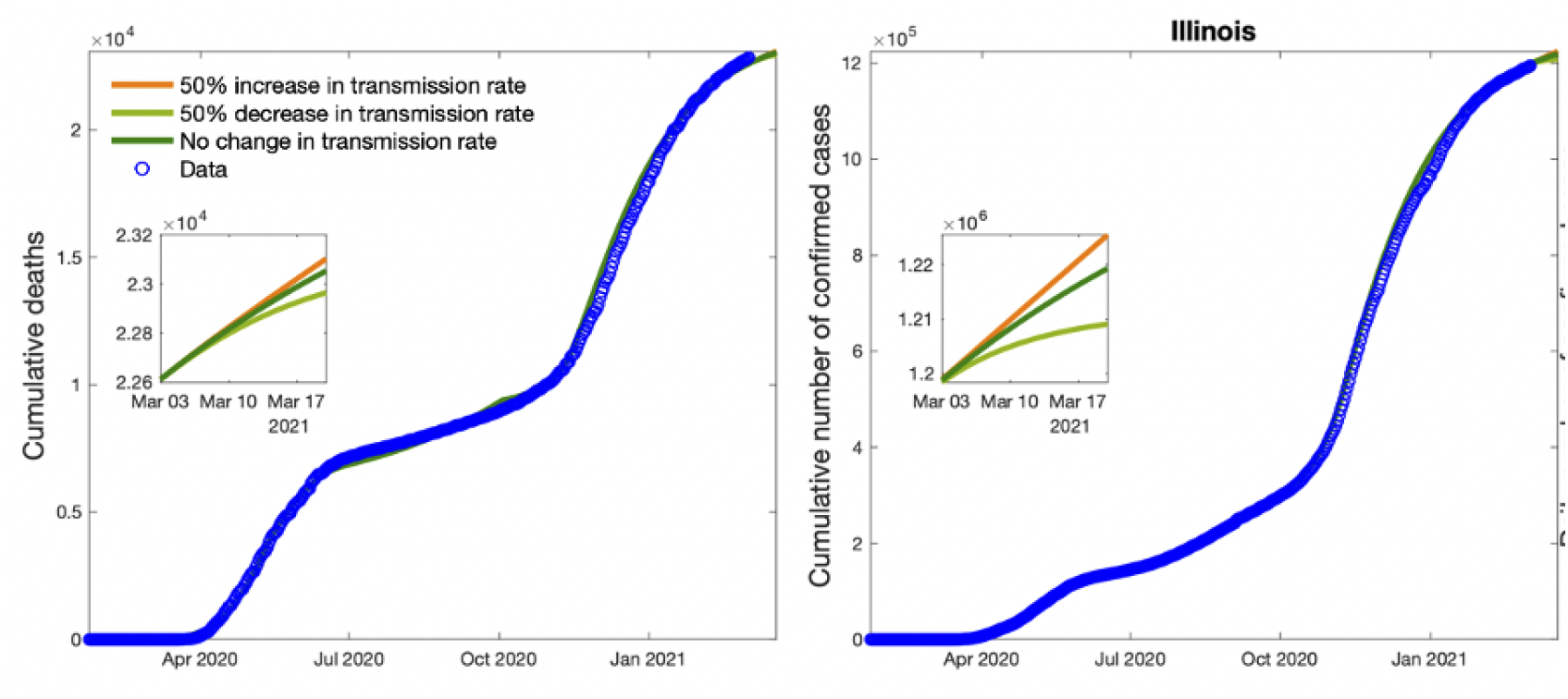
Plots show cumulative cases, deaths, and new cases in Illinois with data. The small boxes in the figures refer to fifteen days forecasting by varying the transmission rate *β* 50% above and below its estimated value.

We see the smallest transmission rate between January 10, 2021 and March 2, 2021. We also forecast the cumulative number of cases and deaths from March 2, 2021 to March 25, 2021. Depending on our 3 scenarios, we forecasted possible number of cases and deaths for Texas (see Table 11).

**Table 11:** Forecasting of the total number of cases, deaths and case fatality rate (CFR) with forecasting up to February 15^th^, 2021 in Illinois, USA

| California | Total # of Cases | Total # of Deaths | CFR |
| --- | --- | --- | --- |
| Scenario 1 | 1,219,250 | 23,052 | 0.019 |
| Scenario 2 | 1,225,390 | 23,101 | 0.019 |
| Scenario 3 | 1,209,040 | 22,963 | 0.019 |

### 4.6 Louisiana

Based on the time series of confirmed cases and deaths from March 2020 to March 2021, the second and third waves are bigger than the first wave (see Figure 8). Depending on the estimated parameters of our model, we see larger transmission rates during the first and third waves (see Table 12). Even if we see a smaller transmission rate in the second wave, the social distancing rate is 0.32, which is much worse than in the first and third wave. The social distancing rates were 0.11 and 0.15 in the first and third waves, respectively (see Table 13). Social distancing rate, *r* causes smallest reduction in the transmission rate between November 13, 2021 and March2, 2021. We also forecast the cumulative number of cases and deaths from March 2, 2021 to March 25, 2021. Depending on our 3 scenarios, we forecasted possible number of cases and deaths for Louisiana (see Table 13).

**Table 12:** Transmission rate, *β*, the reduction rate due to the social distancing, *r*, and the reduced transmission rate due to the social distancing, *rβ* for each corresponding fit in the state of Louisiana

| Fits | $\beta$ | $r$ | $\beta r$ | Date |
| --- | --- | --- | --- | --- |
| First fit | 2.84 | 0.11 | 0.31 | January 25, 2020- May 9, 2020 |
| Second fit | 1.04 | 0.32 | 0.33 | May 10, 2020- June 19, 2020 |
| Third fit | 1.17 | 0.25 | 0.29 | June 20, 2020- October 4, 2020 |
| Fourth fit | 2.23 | 0.15 | 0.34 | October 5, 2020- November 12, 2021 |
| Fifth fit | 1.5 | 0.35 | 0.184 | November 13, 2021- March 2, 2021 |

**Table 13:** Forecasting of the total number of cases, deaths and case fatality rate (CFR) with forecasting up to February 15^th^, 2021 in Louisiana, USA

| California | Total # of Cases | Total # of Deaths | CFR |
| --- | --- | --- | --- |
| Scenario 1 | 450,763 | 9,714 | 0.022 |
| Scenario 2 | 454,118 | 9,741 | 0.022 |
| Scenario 3 | 445,243 | 9,667 | 0.022 |

**Figure 8:**
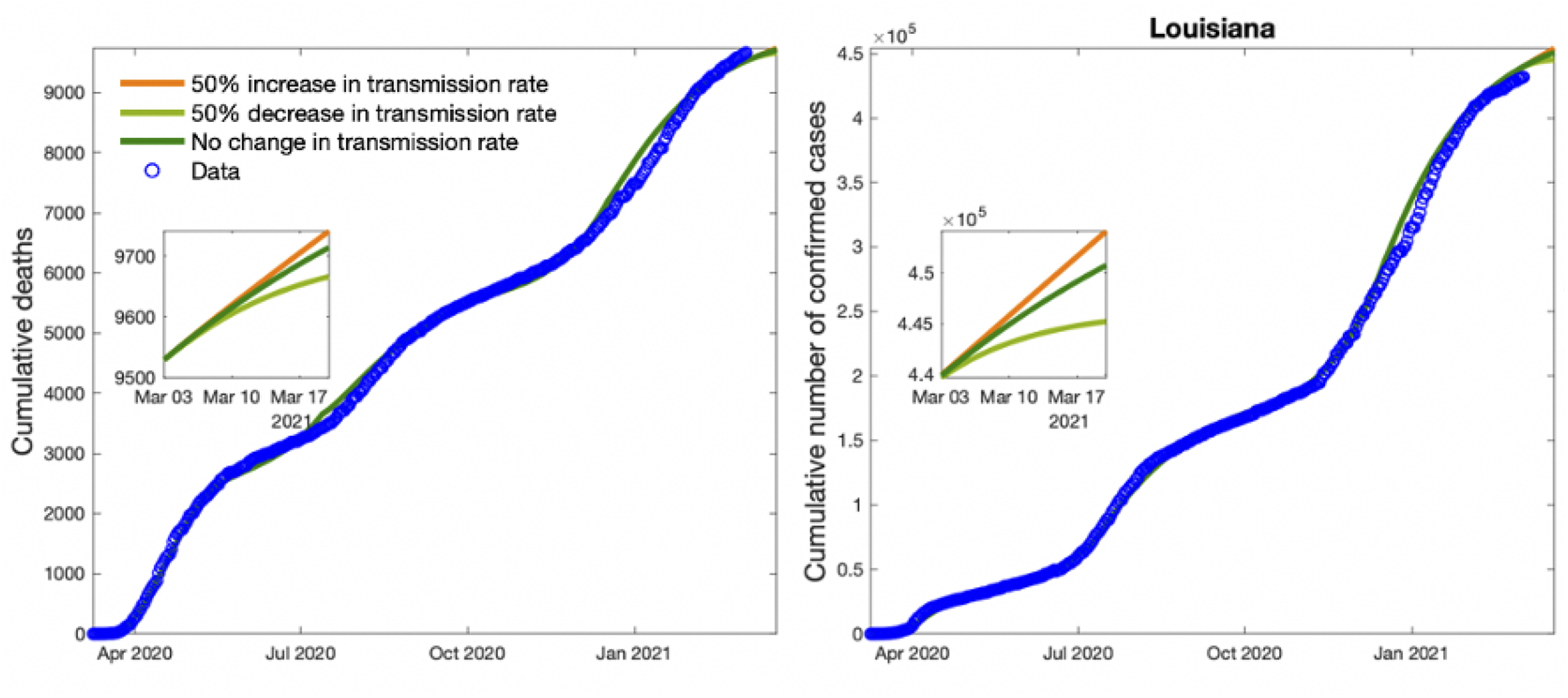
Plots show cumulative cases, deaths, and new cases in Louisiana with data. The small boxes in the figures refer to fifteen days forecasting by varying the transmission rate *β* 50% above and below its estimated value.

### 4.7 Michigan

As it can be seen in Figure 9, we have 3 waves and before the second wave ended the third wave occurred. Based on our estimated parameters, we see larger transmission rates in the first and second wave. Even if we see a smaller transmission rate in the third wave, the social distancing rate is 0.39, which result in a weak social distancing when we compare with the first and second waves. The social distancing rates are 0.1 and 0.17 in the first and second waves, respectively. These rates cause 90% and 83% reduction in transition rates during which (see Table 14). Note that the reduction rate, *r* gets the largest value during October 5th and November 12th which the strongest third wave hits. We also forecast the cumulative number of cases and deaths from March 2, 2021 to March 25, 2021. Depending on our 3 scenarios, we forecasted the possible number of cases and deaths for Michigan (see Table 15).

**Table 14:** Transmission rate, *β*, the reduction rate due to the social distancing, *r*, and the reduced transmission rate due to the social distancing, *rβ* for each corresponding fit in the state of Michigan

| **Fits** | $\beta$ | $r$ | $r\beta$ | **Date** |
| --- | --- | --- | --- | --- |
| First fit | 3.17 | 0.1 | 0.32 | January 25, 2020- May 19, 2020 |
| Second fit | 0.85 | 0.13 | 0.11 | May 20, 2020- July 9, 2020 |
| Third fit | 1.11 | 0.15 | 0.17 | July 10, 2020- September 19, 2020 |
| Fourth fit | 0.56 | 0.39 | 0.22 | September 20, 2020- November 13, 2021 |
| Fifth fit | 0.5 | 0.18 | 0.09 | November 14, 2021- March 2, 2021 |

**Table 15:** Forecasting of the total number of cases, deaths and case fatality rate (CFR) with forecasting up to February 15^th^, 2021 in Michigan, USA

| **California** | **Total # of Cases** | **Total # of Deaths** | **CFR** |
| --- | --- | --- | --- |
| Scenario 1 | 660,105 | 16,605 | 0.025 |
| Scenario 2 | 662,764 | 16,628 | 0.025 |
| Scenario 3 | 655,476 | 16,566 | 0.025 |

**Figure 9:**
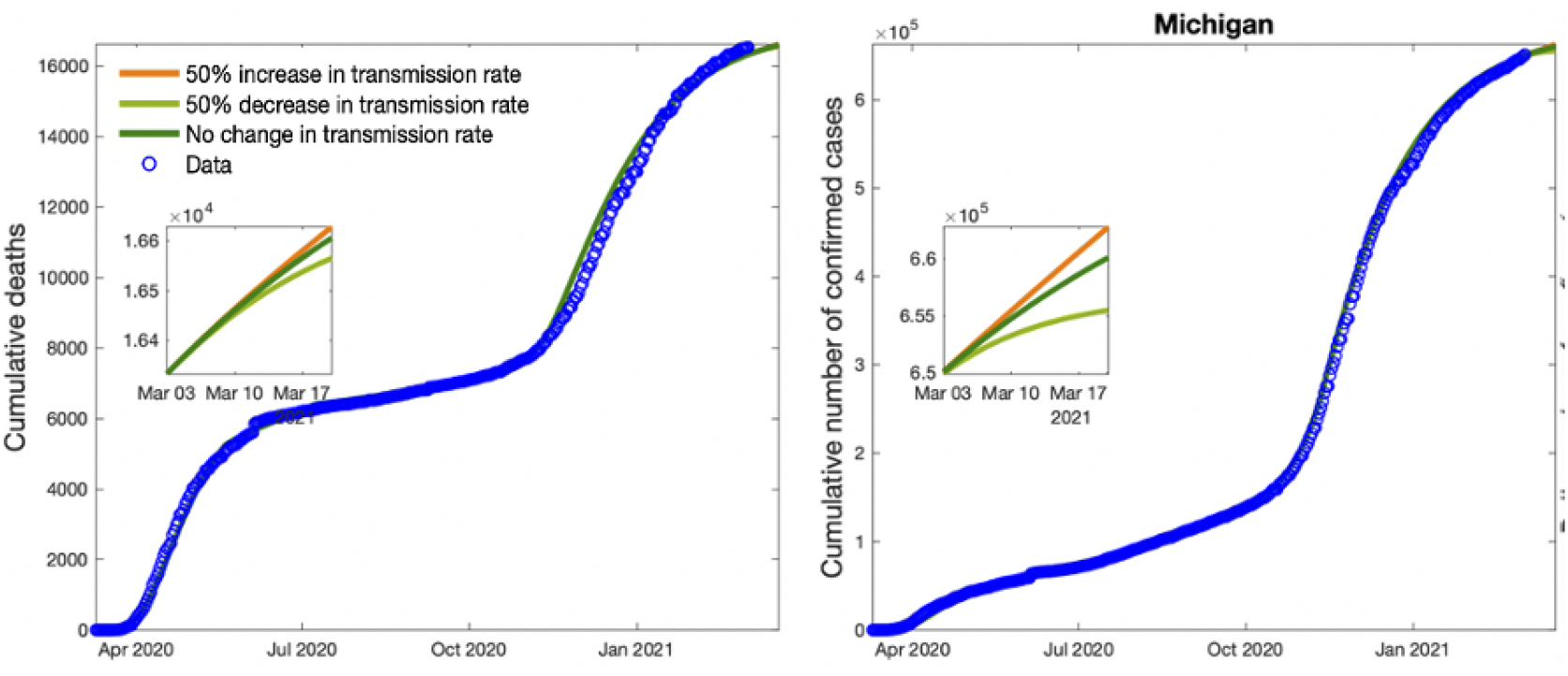
Plots show cumulative cases, deaths, and new cases in Michigan with data. The small boxes in the figures refer to fifteen days forecasting by varying the transmission rate *β* 50% above and below its estimated value.

### 4.8 Missouri

We have 3 waves and the third wave occurred before the second wave was over (see Figure 10). Based on our estimated parameters, we see the larger transmission rates in the first and third waves (see Table 16). Even if the transmission rate of third wave is smaller than transmission rate of the first wave, the reduction in transmission rate of the third wave is about 74 percent (0.26) which is smaller than the reduction of transmission rate of the first wave, which is about 88 percent (0.12). We see the smallest transmission rate between January 10, 2021 and March 2, 2021. We also forecast the cumulative number of cases and deaths from March 2, 2021 to March 25, 2021. Depending on our 3 scenarios, we forecasted possible number of cases and deaths for Michigan (see Table 17).

**Table 16:** Transmission rate, *β*, the reduction rate due to the social distancing, *r*, and the reduced transmission rate due to the social distancing, *rβ* for each corresponding fit in the state of Missouri

| Fits | $\beta$ | $r$ | $r\beta$ | Date |
| --- | --- | --- | --- | --- |
| First fit | 3.24 | 0.12 | 0.38 | January 25, 2020- May 24, 2020 |
| Second fit | 1.06 | 0.28 | 0.3 | May 25, 2020- July 24, 2020 |
| Third fit | 1.2 | 0.24 | 0.29 | July 25, 2020- October 14, 2020 |
| Fourth fit | 2.01 | 0.26 | 0.52 | October 15, 2020- December 6, 2020 |
| Fifth fit | 0.83 | 0.37 | 0.31 | December 7, 2020- March 2, 2021 |

**Table 17:**
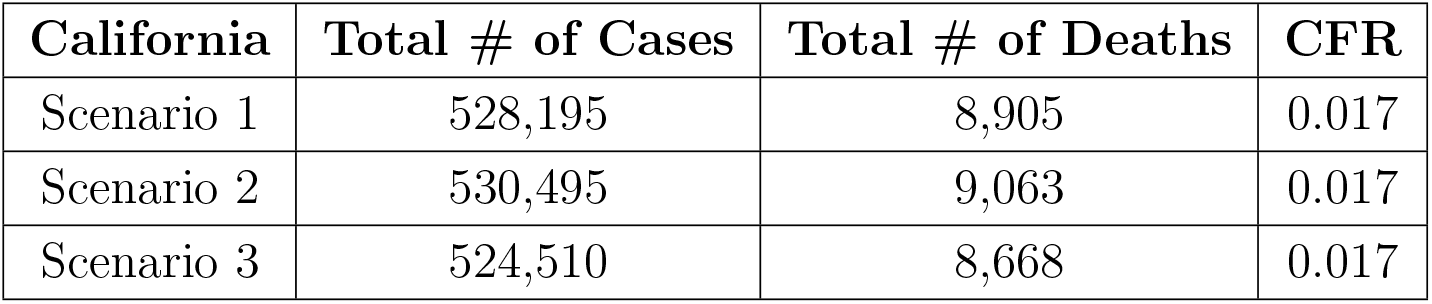
Forecasting of the total number of cases, deaths and case fatality rate (CFR) with forecasting up to February 15^th^, 2021 in Missouri, USA

**Figure 10:**
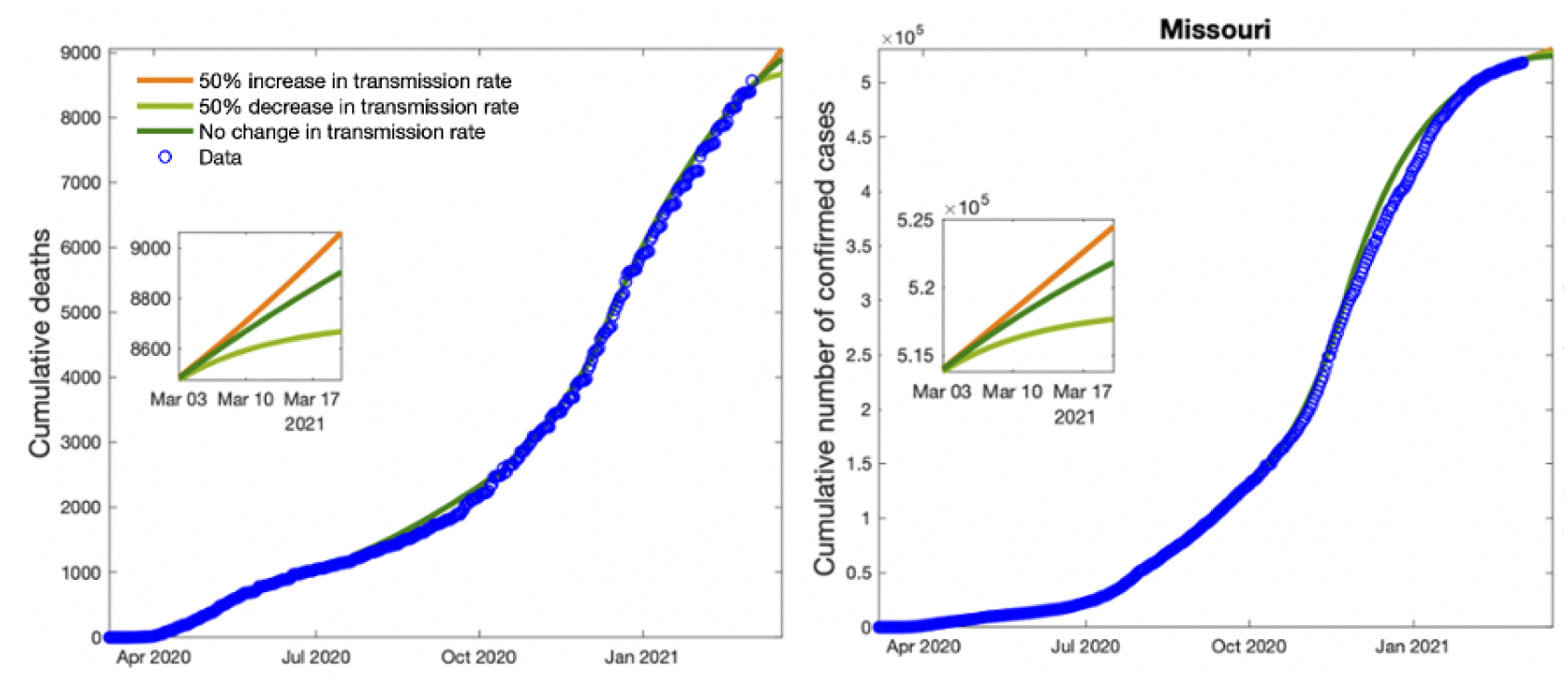
Plots show cumulative cases, deaths, and new cases in Missouri with data. The small boxes in the figures refer to fifteen days forecasting by varying the transmission rate *β* 50% above and below its estimated value.

Our analysis also indicated that the percentage of people in susceptible compartment (people do not social distancing themselves) changes between 10% to 18% from state to state. Thus, about 82 percent to 90 percent people are social distancing themselves (i.e., people wearing mask and avoid social activities) in states we investigate in the study. The amount of people taking social distancing is 90% for California, 89% for Texas, 84% for Florida, 82% for Georgia, 88% for Illinois, 89% for Louisiana, 89% for Michigan, and 90% for Missouri. Note that we just considered staying at home order between January, 2020 and May, 2020 by using the model 1 in Eq. (1). After May, 2020, we considered *S*_*d*_ class as the social distancing class instead of considering it as the self-quarantine (staying at home) class by using the model 2 in Eq. (2). As discussed in the section 2.1, individuals in the *S*_*d*_ compartment are just practicing social distancing rules mandated by government authority.

Social distancing compartment (*S*_*d*_), reduces transmission rate of COVID-19 between 60% and 90% for California, between 76% and 88% for Texas, between 79% and 87% for Florida, between 63% and 90% for Georgia, between 60% and 90% for Illinois, between 65% and 89% for Louisiana, and between 61% and 90% for Michigan,and between 63% and 88% for Missouri between January 2020 and March 2021 (see the reduction rate due to taking social distancing, *r* in Tables for each corresponding state).

### 4.9 Conclusion and Discussion

Practicing social distancing (i.e., using mask, avoiding contact with others, and obeying all the rules recommended by state governments) reduces transmission rates between 60% and 90% for the states. However, we see an increase in the reduction rate due to the social distancing from January 2021 to March 2021, which indicates the social distancing level was decreasing over time.

Our study also indicated that the percentage of people who is not taking social distancing changes between 10 percentage to 18 percentage from state to state. The states like Missouri and Illinois have a larger percent of people practicing social distancing, so any increases in the reduced transmission rate, *rβ* results in larger wave comparing to the states that have a lower percentage of people practicing social distancing like Georgia and Florida.

Even though some states have similar reduced transmission rates, *rβ*, we see stronger waves in the states, which have a larger reduction rate, *r*. For example, see the tables and figures of California and Illinois. This indicates the reduction rate, *r* plays a crucial role to reduce the magnitude of the waves.

Based on our analysis, without further precautions (i.e., more strict restrictions or efficient vaccination), it does not look possible to contain the outbreak with current strategies taken in the states.

## Data Availability

World Health Organization
New York Times

https://www.who.int/

https://github.com/nytimes/covid-19-data

